# Whole Genome Landscape Analysis of Homologous Recombination Deficiency in a Pan-Cancer Cohort

**DOI:** 10.1101/2024.06.28.24309592

**Authors:** Majd Al Assaad, Kevin Hadi, Max F. Levine, Daniela Guevara, Minal Patel, Marvel Tranquille, Abigail King, John Otilano, Alissa Semaan, Gunes Gundem, Juan S. Medina-Martínez, Michael Sigouros, Jyothi Manohar, Hui-Hsuan Kuo, David C. Wilkes, Eleni Andreopoulou, Eloise Chapman-Davis, Scott T. Tagawa, Andrea Sboner, Allyson J. Ocean, Manish Shah, Elli Papaemmanuil, Cora N. Sternberg, Kevin Holcomb, David M. Nanus, Olivier Elemento, Juan Miguel Mosquera

## Abstract

**Purpose:** Homologous recombination deficiency (HRD) impacts cancer treatment strategies, particularly the effectiveness of PARP inhibitors. However, the variability different HRD assays has hampered the selection of oncology patients who may benefit from these therapies. Our study aims to assess the whole genome landscape to better define HRD in a pan-cancer cohort and to contribute to harmonization of HRD detection.

**Methods:** We employed a whole-genome sequencing WGS HRD classifier that included genome-wide features associated with HRD to analyze 580 tumor/normal paired pan-cancer samples. The HRD results were correlated retrospectively with treatment responses and were compared with commercial HRD tests in a subset of cases.

**Results:** HRD phenotype was identified in 62 samples across various cancers including breast (19%), pancreaticobiliary (17%), gynecological (15%), prostate (8%), upper gastrointestinal (GI) (2%), and other cancers (1%). HRD cases were not confined to *BRCA1/2* mutations; 24% of HRD cases were *BRCA1/2* wild-type. A diverse range of HRR pathway gene alterations involved in HRD were elucidated, including biallelic mutations in *FANCF, XRCC2*, and *FANCC*, and deleterious structural variants. Comparison with results from commercial HRD assays suggests a better performance of WGS to detect HRD, based on treatment response.

**Conclusion:** HRD is a biomarker used to determine which cancer patients would benefit from PARPi and platinum-based chemotherapy. However, a lack of harmonization of tests to determine HRD status makes it challenging to interpret their results. Our study highlights the use of comprehensive WGS analysis to predict HRD in a pan-cancer cohort, elucidates new genomic mechanisms associated with HRD, and enables an accurate identification of this phenotype, paving the way for improved outcomes in oncology care.

## Introduction

*BRCA1* and *BRCA2* (*BRCA1/2*) play a significant role in an error-free DNA damage repair pathway known as homologous recombination repair (HRR) ^1^. This pathway corrects DNA double-strand breaks (DSBs) and interstrand cross-links ^1,2^. Somatic or germline mutations in *BRCA1/2* (*BRCA*mut) lead to homologous recombination deficiency (HRD) and have long been associated with breast, ovarian, pancreatic, and prostate cancers ^3,4^. Poly (ADP-ribose) polymerase inhibitors (PARPi) have been developed to treat cancers associated with mutations in *BRCA1/2* and other HRR genes based on the synthetically lethal relationship with HRD ^5^. Furthermore, the use of alkylating-like agents, especially platinum-based chemotherapy (PBCT), has shown increased effectiveness for cancers with HRD ^6,7^.

Loss of function in HRR genes, such as *RAD51B, ATM, FANC* genes, *CHEK2, PALB2*, among others, can result from small mutations, structural variants, or epigenetic changes ^8-10^. Therefore, evaluation of different deleterious mechanisms would require several assays and advanced analysis. Due to the broad range of HR-inactivating genes, it is necessary to not only survey the full genetic landscape of HR genes, but also detect DNA damage signatures associated with HRD for selective treatment with PARPi and PBCT ^10^.

HRD is associated with DNA damage signatures, including single base substitution (SBS), structural variant (SV) signatures, in addition to loss of heterozygosity (LOH), large scale transition (LST), and telomeric allelic imbalance (TAI) ^11-13^. Assays that report HRD, such as MyChoice® CDx and FoundationOne CDx, among others, are based on targeted next-generation sequencing (NGS) and only employ a subset (LOH, LST, and TAI) of the available signals to detect HRD. Their use in PARPi treatment of ovarian cancer has been approved based on the results of multiple clinical trials ^14-17^. It is worth noting that in some trials several patients with *BRCA*ness have experienced shorter survival under first-line maintenance PARPi and numerous cases lacking *BRCA*ness have shown extended survival (16, 20). In contrast to the current commercially available companion diagnostics (CDx) for HRD testing, algorithms using whole genome sequencing (WGS) such as HRDetect and CHORD, employ all mutation classes. However, these have yet to be tested in clinical settings.

The clinical and research assays have shown a variable association between *BRCA*ness and the presence of HRR genes mutations ^3,18,19^. This variability indicates that not every deleterious mutation in these genes results in *BRCA*ness. Conversely, *BRCA*ness can also be associated with *BRCA1/2* wild type (*BRCA*wt) and variants of unknown significance (VUS) ^18^. This highlights the necessity of focusing on the *BRCA*ness phenotype rather than solely relying on the known pathogenicity of mutations.

WGS offers a comprehensive assessment of the genome, covering simple and complex structural variants, copy number alterations, and mutational patterns associated with HRD ^20,21^. It achieves superior precision and sensitivity in identifying *BRCA*ness. Consequently, WGS-based HRD biomarkers may more effectively stratify cancer patients for therapeutic interventions ^22^.

In this study we analyzed WGS data from 580 pan cancer samples, investigated the prevalence and characteristics of HRD employing a novel WGS based classifier, compared the WGS-based HRD results with scores from commercial panels, and correlated them with treatment responses.

## Methods

### I. Patient enrollment, tissue samples and clinical data acquisition

Participants were prospectively enrolled at Weill Cornell Medicine in the Institutional Review Board (IRB)-approved protocols WCM IRB #1305013903 and #1007011157. Tumor DNA for WGS was extracted from frozen tumor samples, formalin-fixed, paraffin-embedded (FFPE) archival tissue, or fluid from malignant ascites. Histopathology review was performed before DNA extraction. Germline DNA was primarily extracted from blood. When unavailable, saliva or benign tissue (frozen or FFPE) was used. We collected comprehensive clinical data, which encompassed age, sex, ethnicity, treatment-related information, radiologic findings, and pathologic data. Sequential study IDs were assigned to the samples using the format “WCM” followed by a number. These IDs are known only within the research group.

### II. DNA Extraction and WGS

For DNA extraction from FFPE blocks, we used 5-micrometer-thick unstained slides macrodissected for at least 80% tumor content. For DNA extraction from frozen specimens, we employed 3mm core punches from the frozen OCT-embedded tissue. The Maxwell® 16 FFPE Plus DNA kit (Promega, Cat# AS1135) was employed, in combination with the Maxwell® 16 instrument (Promega, Madison, WI). DNA quality and quantity was assessed by using the Agilent Tapestation 4200 (Agilent Technologies) and the Qubit Fluorometer (ThermoFisher), respectively. Whole genome sequencing (WGS) was carried out at the New York Genome Center on an Illumina Novaseq6000 sequencer using 2x150 bp cycles. Libraries were generated using the KAPA Hyper Library Preparation Kit (KAPABiosystems KK8502, KK8504), targeting 500 bp fragments, in compliance with the manufacturer’s instructions. DNA fragments underwent a series of preparation steps including shearing, end-repair, adenylation, and ligation to Illumina sequencing adapters. The prepared DNA fragments were size-selected using bead-based methods and amplified ^23^. Quality and quantity of the final libraries were assessed prior to sequencing.

### III. WGS data processing pipeline and HRD curation

We employed the Isabl GxT analytics platform to process all WGS and RNA sequencing data and generate comprehensive reports ^24^. Within the Isabl pipeline deployed in AWS cloud HPC environments, DNA and RNA alignment with BWAMem and STAR, quality control, somatic and germline variant calling, and annotation were performed as previously described ^25-29^. Briefly, ensemble calling was done for both somatic and germline single nucleotide variants (SNV), InDels, and somatic SVs. These variant classes were then annotated and those in which at least 2 of 3 callers of each class were included for reporting. Purity, ploidy, and genome-wide copy number states were estimated with Battenberg, followed by annotation of CNA events at the gene level. Driver alterations for all variant classes and their potential treatment targets were assessed by cross-referencing protein-coding variants with COSMIC and OncoKB.

WGS-HRD scores were assigned with Isabl HRD, random forest classifier trained on a subset of the data (N=321 patients) and described in and Hadi et al. (abstract) ^30^ to detect HRD by incorporating evidence from genome-wide SNV, indels, SV, and CNV signals (**Figure S1**). Briefly, individual patients with HRD were identified as the top quartile outliers for features associated with HRD (COSMIC SBS3, deletions with microhomology, and small SV duplications and deletions). Cases that had at least two outlier signatures were manually inspected for presence of HRD by visualizing circos plots and signature frequencies to identify those with HRD. 96 trinucleotide SNV contexts, 45 InDel types, 38 SV types, and LST, LOH, and TAI scores were used as features for the classifier. On a validation cohort of 556 PCAWG samples, the classifier achieved a high AUROC/AUPRC of 0.99/0.96 ^30^. We applied the classifier on the entire cohort, which assigned a probability of presence of the HRD phenotype. The resulting score ranged from 0 to 1, with a score equal to or greater than 0.5 indicating HRD (≥0.5) and a score less than 0.5 labeled as HRR proficient (HRP <0.5). Tumor purity > 20% is required to accurately assess HRD.

### IV. Fluorescence in situ Hybridization

Four-µm-thick formalin-fixed paraffin-embedded tissue sections were used for fluorescence in situ hybridization (FISH) analysis, as described in our protocols ^31-33^. Bacterial artificial chromosomes were designed against loci of interest to prepare break-part dual-color FISH probes ^34^. For *BRCA*2 RP11-110O22 BAC clone was labeled red and RP11-11K16 clone was labeled green, and for *ATM* RP11-144G7 BAC clone was labeled red and RP11-589O5 clone was labeled green. All clones were validated on normal metaphase spreads before any application on FFPE tissue. A positive break-apart was determined by one red, one green, and one yellow signal (combination of red and green signal indicating the normal chromosome homologue). At least 200 nuclei were analyzed per case using a fluorescent microscope (Olympus BX51; Olympus Optical). Cytovision 7.3.1 software was used for imaging and analysis.

## Results

### Sample Characteristics and Frequency of HRD

We performed WGS analysis on 580 samples from 453 patients. **Figure 1** summarizes the types of tumor samples (FFPE, frozen, or fluid) and type of lesion (primary and metastatic). These encompass 77 unique histology types (**Figure S2**), classified according to the MSKCC Oncotree ^35^, and obtained from 33 unique primary sites. Sites were divided into six cancer subgroups: prostate (29%), gynecological (20%), pancreaticobiliary (15%), breast (11%), upper gastrointestinal (GI) (10%), and “others” (12%), which included smaller cancer cohorts and rare tumors (**Figure 1, Figure S2**).

**Fig 1.**
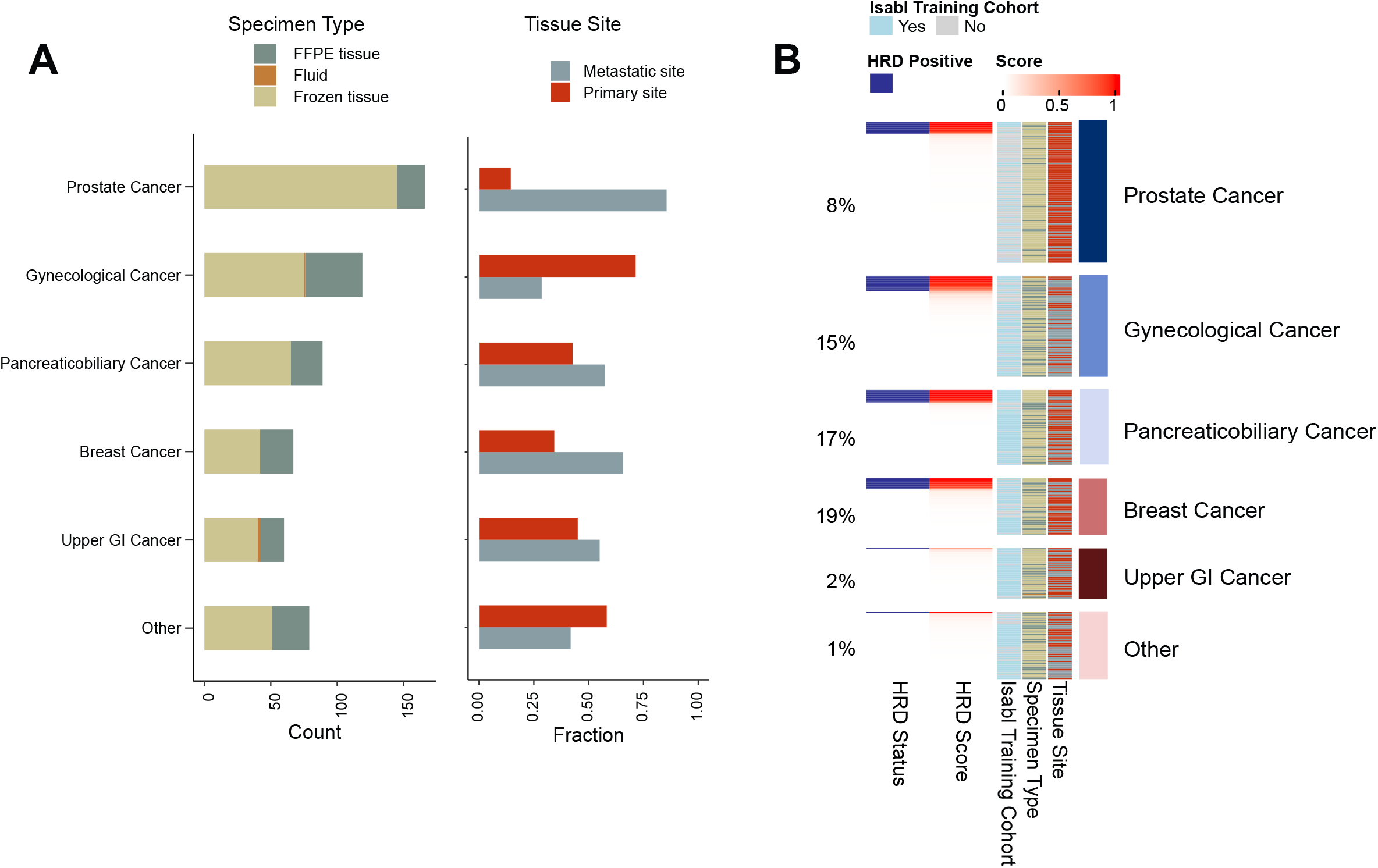
Cohort Characteristics. (A) Left panel, overall breakdown of cohort by specimen type (left) and whether biopsy was from a primary or metastatic tumor (right). (B) Breakdown of the Isabl HRD score and HRD status (cutoff > 0.5) across the cohort. FFPE, formalin-fixed, paraffin-embedded; GI, gastrointestinal; HRD, homologous recombination deficiency.

Among the total 580 samples, 62 samples across 53 patients exhibited HRD by WGS. The highest percentage of HRD cases were found in breast cancer (19%) followed by pancreaticobiliary (17%), gynecological (15%) and prostate cancers (8%). Upper GI cancers had the lowest percentage of HRD with only one case. In addition, 1 case of carcinoma of unknown primary in the “others” cohort harbored HRD.

### Mutational landscape of HRD

While *BRCA*ness is most commonly explained and clinically tested by alterations in *BRCA1/2*, dysfunction in other HRR pathway genes can also result in phenocopy. We investigated events in *BRCA1*, *BRCA2*, and other HRR pathway genes in HRD cases, accounting for loss of heterozygosity and compound hits that could result in biallelic loss of function and focusing on SVs and somatic mutations with unknown effects on HRD (**Figure 2.A**). Out of 62 HRD samples across 53 patients, 76% (47) harbored alterations in *BRCA1* and/or *BRCA2*(*BRCA1/2mut*). In 55% (34) of samples, *BRCA1/2mut* had biallelic pathogenic small mutations (SNVs and InDels) and 6% (4) had homozygous deletions. Another 3% (2) harbored SVs with LOH in *BRCA1/2* with predicted impact to coding sequence. Interestingly, none of those two cases with *BRCA1/2* biallelic SVs had deleterious mutations in other HRR pathway-related genes (**Figure 2**). 11% (7) harbored *BRCA1/2* small mutation VUS or SV VUS. Ultimately, the remaining 15 HRD cases (24%) were *BRCA*wt (**Figure 2.A and Figure 2.B**).

**Fig 2.**
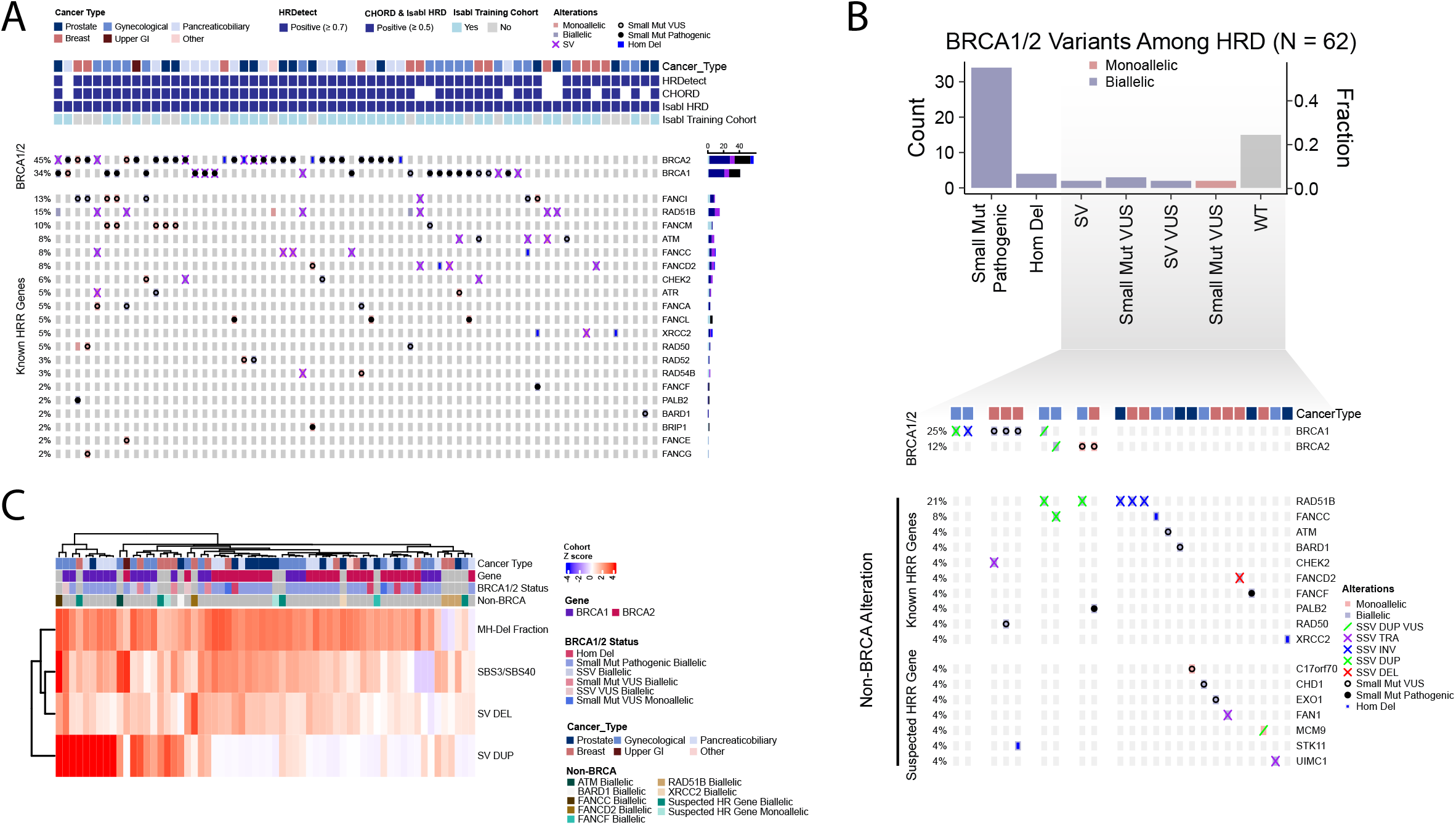
Genomic alterations and features of HRD. (A) Landscape of *BRCA1/2* and other HR-associated genes in HRD cases. (B) Top, *BRCA1/2* alteration status in the HRD cohort by variant class. Bottom, further delineating alterations in genes outside of *BRCA1/2* in cases that have *BRCA1/2* events not considered in standard clinical practice (SSV or VUS) or are *BRCA*wt. (C) Genomic feature enrichment (Z-score calculated across the full HRD & HRP cohort) in HRD. Notably, the features of HRD are not distinguished by gene target or variant type. Del, deletion; Dup, duplication; GI, gastrointestinal; Hom, homozygous; HRD, homologous recombination deficiency; HRR, homologous recombination repair; MH, microhomology; Mut, mutation; SBS, single base substitution; SV, structural variant; VUS, variant of unknown significance; WGS, whole genome sequencing; WT, wild-type.

For the 15 HRD samples with *BRCA*wt we investigated other gene mutations that might have caused the HRD phenotype and we found that 1 had a pathogenic small mutation in *FANCF*, 1 had biallelic deletion in *XRCC2*, and 1 had biallelic *FANCC* deletion. The remaining *BRCAwt* HRD cases (n=12) either had no mutations in any of the known HRR genes (n=5; 33% of the *BRCA*wt HRD cases) or had a VUS small mutation or SV in one or multiple HRR pathway genes including *RAD51B, PALB2, ATM* (**Supplementary figure 2**), *CHEK2, FANC*D2 and *RAD51* (n=7; 47% of the *BRCA*wt HRD cases).

Features frequently enriched in the HRD cohort consisted of genome wide small microhomology deletions(MH-dels), COSMIC signatures (V3) SBS3 or SBS40, SV deletions from 1-10kb, and SV duplications from 1-10kb (**Figure 2.C**). SV duplications were largely enriched in *BRCA*1 mutated tumors. While MH-dels were the most frequently enriched feature among HRD samples, 3 *BRCA*wt, *RAD51B* mutated samples harbored intermediate MH-dels with enrichment of SV deletions or duplications and SBS3/40 indicating a potentially distinct HRD feature profile.

It is worth noting that 9 out of 62 HRD samples represented additional biopsies. These 9 harbored similar *BRCA1/2* status as the original patient sample (8 small pathogenic *BRCA*mut, 1 *BRCA*wt). In this series of patients, HRD was consistently present across all except 1 patient in which the primary tumor (HRD score 0.94) harbored an SV breakpoint in exon1 of *BRCA*1 coinciding with LOH while the metastasis was borderline (0.44) and did not harbor evidence of the *BRCA*1 SV nor LOH.

We also investigated HRD positivity using two other WGS based HRD algorithms, CHORD and HRDetect (**Figure 2.A**) ^3,19^. Overall, 9 HRD samples were negative by CHORD, which included 3 cases that are simultaneously negative by HRDetect. Interestingly, 2 of those cases were *BRCA*wt with *RAD51* structural variants, and one with *BRCA*2 monoallelic VUS somatic small mutation and *BRCA*1 biallelic pathogenic somatic small mutation.

### Clinical and genomic characteristics of HRD cases with *BRCA*wt and *BRCA1/2* SV

Standard clinical decisions to treat HRD cancers largely relies on the presence of small mutations or copy alteration in *BRCA1/2* based on targeted panel testing. Having analyzed the mutational and feature landscape of HRD on our cohort using WGS, we then sought to examine the patient’s response to treatment in HRD cases in which clinical history was available and which presented no *BRCA1/2* small mutations or copy alterations.

The first case, WCM414, is a patient diagnosed with high-grade serous fallopian tube carcinoma (HGSFC). She underwent resection then PBCT. WGS demonstrated HRD, indicated by the presence of tandem duplications and 56% contribution from SBS3. Additionally, she had *BRCA*wt with a biallelic small VUS mutation in *ATM*. The corresponding MyChoice CDx test also indicated HRD and confirmed *BRCA*wt status. Consequently, she was administered maintenance PARPi and has shown no evidence of disease recurrence to date, with a PFS of 20 months.

The second case, WCM209, is a patient with high-grade serous carcinoma of the ovary (HGSOC) (**Figure 3.A**). WGS analysis showed no mutations in *BRCA*1, *BRCA*2, or other HRR pathway genes, a finding that is consistent with the patient’s clinical genomic testing, which included FoundationOne and Invitae germline mutation assays. Notably, WGS indicated HRD, characterized by a SBS3 signature contribution of 22% and an increase in small deletions and tandem duplications (**Figure 3.A**). A relevant finding was a translocation resulting in a *HIF1A*::*UIMC1* fusion involving intron 13 of *UIMC1*, coupled with LOH. *UIMC1* (Ubiquitin Interaction Motif Containing 1) encodes a nuclear protein that collaborates with *BRCA*1 in recognizing and repairing DNA lesions ^36^. This fusion was validated by RNA sequencing analysis (**Figure 3.A**). MyChoice CDx testing was also performed, which indicated HRD. The patient was treated with PBCT, achieving near-complete resolution of the metastatic tumor, as assessed by RECIST 1.1 criteria.

**Fig 3.**
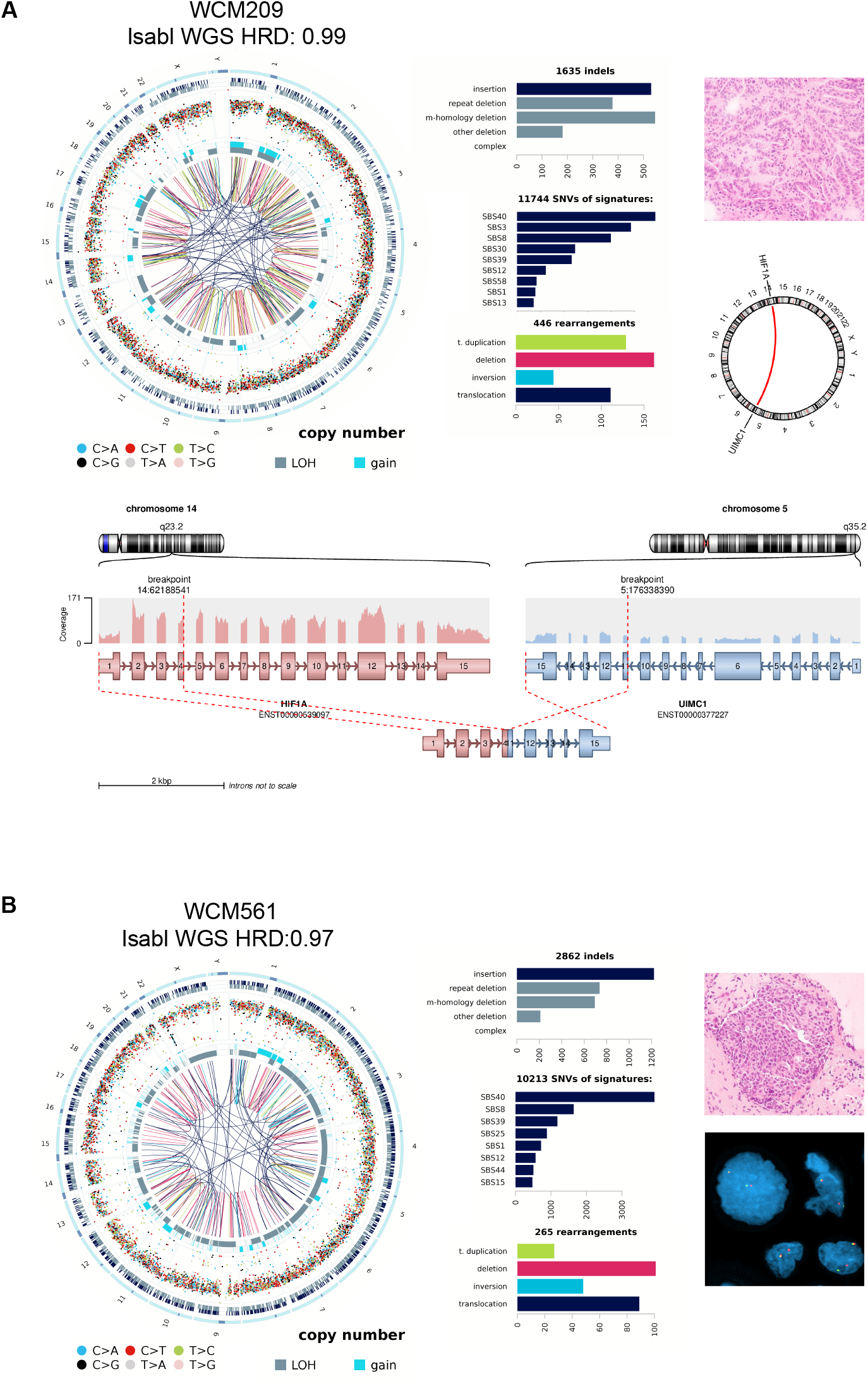
Validation of alterations in 2 WGS HRD positive cases. (A) HGSOC (H&E, 20x) tumor harboring HRD signals as shown in the circos plot with high rate of microhomology deletions, SBS3, SV tandem duplications, and SV deletions. WGS detected an in-frame *HIF1A*::*UIMC1* fusion that leads to a truncated, non-functional chimeric transcript that is missing exon 6, necessary for binding to *BRCA*1. (B) PRNE (H&E, 20x) tumor with HRD that harbored a *BRCA*2::*TMPRSS2* fusion validated by FISH. H&E, hematoxylin and eosin; HGSOC, high grade serous ovarian carcinoma; HRD, homologous recombination deficiency; Indel, insertion deletion; LOH, loss of heterozygosity; PRNE, prostate neuroendocrine carcinoma; SBS, single base substitution; SV, structural variant.

The third case (WCM561) is a patient diagnosed with advanced prostatic carcinoma. WGS was performed on a metastatic brain lesion that exhibited a neuroendocrine histology (**Figure 3.B**). WGS revealed a partial homozygous loss in *BRCA2* that was caused by an SV translocation at intron 21 resulting in a *BRCA2*::*TMPRSS2* fusion accompanied by LOH in the intact portion of the gene. The presence of this fusion was validated by FISH, which demonstrated *BRCA2* break-apart signals. WGS also indicated HRD in this case, with increased small deletions (**Figure 3.B**). Unfortunately, treatment response data for this patient was not available.

These examples illustrate how WGS can identify additional HRD cases, particularly those with *BRCA*wt.

### Comparison of WGS HRD with commercially available assays

Current commercial tests for HRD employ only GIS or LOH scores based on allele-specific copy number segmentation, a subset of genome-wide features predictive of HRD and captured by WGS. To explore the potential clinical significance of WGS-based HRD testing, we retrospectively assessed cases with WGS, commercial HRD scores, and clinical history that included treatment response. From the 39 serous carcinoma cases in the gynecological cancer cohort, where an HRD score can inform treatment decisions, we identified 16 cases that had undergone a commercial assay that included an HRD score. This group comprised 9 cases evaluated with MyChoice CDx GIS and 7 cases with FoundationOne HRD score. The results of both commercial assays and WGS testing are summarized in **Table 1**. Of these cases, 3 out of 5 identified as HRD-positive by WGS were deemed HRP by commercial assays; these included 1 case tested by FoundationOne (negative) and 2 cases by MyChoice CDx, one negative and the other inconclusive for HRD. Among cases determined as HRP by WGS, 3 out of 10 were found to be positive according to FoundationOne. To delve deeper into the reasons for these discrepancies and assess their potential clinical significance, we analyzed the genomic features of all cases with divergent results. We also retrospectively correlated the scores with responses to PBCT and PARPi treatments, as per RECIST 1.1 criteria, where available.

**Table 1.** Comparison of WGS HRD with commercially available assays.

| WCMID | Isabl | Commercial Assay | Result | Clinical response to Maintenance PARPi |
| --- | --- | --- | --- | --- |
| WCM231 | HRD | FoundationOne | HRP | Progression 10 months after maintenance therapy (PFS = 10months) |
| WCM209 | HRD | MyChoice CDx | HRD | No available PARPi response data |
| WCM414 | HRD | MyChoice CDx | HRD | No disease recurrence to date (PFS= 20 months) |
| WCM205 | HRD | MyChoice CDx | HRP | Did not receive PARPi |
| WCM228 | HRD | MyChoice CDx | Inconclusive | No disease recurrence to date (PFS= 60 months) |
| WCM425 | HRP | FoundationOne CDx | HRD | Progression 2 months after maintenance therapy (PFS = 2 months) |
| WCM295 | HRP | MyChoice CDx | HRP | Did not receive PARPi |
| WCM436 | HRP | FoundationOne CDx | HRD | Did not receive PARPi |
| WCM236 | HRP | FoundationOne CDx | HRD | Progression 3 months after maintenance therapy (PFS = 3 months) |
| WCM435 | HRP | MyChoice CDx | HRP | Did not receive PARPi |
| WCM442 | HRP | FoundationOne CDx | HRP | Did not receive PARPi |
| WCM272 | HRP | FoundationOne CDx | Inconclusive | Progression 3 months after maintenance therapy (PFS = 3 months) |
| WCM284 | HRP | MyChoice CDx | HRP | Did not receive PARPi |
| WCM331 | HRP | MyChoice CDx | HRP | Did not receive PARPi |
| WCM419 | HRP | MyChoice CDx | HRP | Did not receive PARPi |

### Cases with commercial testing negative and WGS homologous recombination deficiency (HRD) status

WCM231 sample was obtained from a HGSOC peritoneal metastases. FoundationOne testing indicated HRD-negative status, although it revealed a pathogenic *BRCA*1 E111fs*1 mutation. WGS confirmed the same *BRCA*1 pathogenic mutation with LOH and showed a HRD phenotype with a high fraction of microhomology deletions (56%) with a high prevalence of COSMIC SBS3 (>6000), and small SV deletions (50) and duplications (30). The patient was treated with neoadjuvant PBCT and exhibited a partial response to treatment according to RECIST 1.1 criteria. The patient was maintained on PARPi for 10 months before experiencing recurrence.

WCM205 sample was obtained from a HGSOC splenic metastasis. MyChoice CDx showed HRP and *BRCA*wt. WGS did not detect *BRCA*mut and other HRR pathway genes mutations. However, WGS showed an HRD phenotype with a moderate high fraction of microhomology deletions (34%) and enrichment of SBS3 (>3000) with a high load of small SV deletions (49). Treatment with PBCT for recurrent peritoneal carcinomatosis showed partial response according to RECIST 1.1 criteria.

Lastly, WCM228 HGSOC sample was obtained from the ovary. MyChoice CDx testing yielded inconclusive results for HRD. Targeted NGS testing identified a *BRCA*2 VUS. WGS confirmed the same monoallelic *BRCA*2 VUS mutation and revealed a *RAD51B* intron 7-intron 8 deletion with loss of heterozygosity. WGS also showed an HRD phenotype with a high fraction of microhomology deletions (49%), high load of SV deletions (31) and duplications (30), and a moderate load of SBS3 (>3000). RECIST 1.1 assessment of the response to the neoadjuvant PBCT showed an overall partial response to treatment. The patient was later maintained on rucaparib and did not experience disease recurrence for 5 years following the diagnosis.

### Cases with Commercial testing positive and WGS homologous recombination repair-proficient (HRP) status

WCM425 sample originated from the HGSOC omental metastasis. FoundationOne testing indicated HRD-positivity with *BRCA*wt, and it identified an *APC* VUS. WGS revealed a HRP phenotype. Rucaparib, used for maintenance therapy, was discontinued two months later due to disease progression.

WCM436 was obtained from another HGSOC omental metastasis. FoundationOne showed HRD with *EP300* mutation and *BRCA*wt. WGS showed HRP. However, tumor purity as determined by WGS was low (0.12), and cannot derive a confident result. The RECIST 1.1 assessment showed a response to treatment with PBCT.

WCM236 sample was a HGSOC primary. FoundationOne testing indicated HRD. WGS showed an HRP phenotype, although it identified a biallelic *RAD51B* translocation at exon 14. The patient showed disease progression 3 months after starting olaparib maintenance (RECIST 1.1).

## Discussion

The significance of HRD in precision oncology has grown because of the development of PARPi and the correlation with response to PBCT in solid tumors ^20,37-40^. Targeted NGS panels are limited in detecting the full range of *BRCA1/2* mutational events and mainly focus on exonic regions ^41-43^. Targeted panels may also overlook alterations in other genes of the HRR pathway that may cause HRD ^41-43^. Consequently, recent research has focused on scoring the genomic signatures of the disruption in the HRR pathway like LOH, TAI, LST, SBS3, small deletions, and tandem duplications ^1,21,43^. However, these HRD ‘footprints’ are not fully revealed by common targeted panels and exome sequencing ^1,44^. To address these limitations, we employed a WGS approach to better determine the HRD status in 580 tumor/normal paired samples and correlated the results with clinical outcomes.

Epithelial carcinomas of the ovary and fallopian tube are the only cancer types approved for PARPi treatment based on clinical HRD signature testing, according to the most recent NCCN guidelines ^45^. In our cohort, we compared WGS HRD phenotype testing results to MyChoice CDx and FoundationOne CDx when available. We identified several discrepant cases: patients with HRD on WGS but HRP by clinical panels showed treatment response to PARPi and/or PBCT, whereas those patients with HRP tumors by our WGS classifier but HRD on other panels exhibited treatment resistance to PARPi or PBCT. A WGS classifier of HRD might reduce false negatives and increase the inclusion of appropriate patients in relevant clinical trials ^46^.

In an effort to assess concordance across commercial and research tests that employ GIS or LOH scores, recent analysis of 13 GIS-based assays revealed challenges in unifying a definition of HRD and reported a wide range of percent positive HRD cases (>50% differences) in the cohorts studied ^47^. More fundamentally, comparative studies that continue to be designed around performance metrics other than treatment or other clinical response limit objective assessment of the use of such assays. While preliminary and limited by the number of cases, our study provides clinical evidence that WGS would improve prediction of HRD with subsequent treatment response, beyond current FDA approved CDx for HRD.

PARPi are progressively becoming a part of the therapeutic arsenal against tumors with HRD. However, treatment of non-ovarian cancers with PARPi depends on specific conditions, according to NCCN guidelines. These include the presence of a pathogenic *BRCA1/2* mutation for breast and pancreatic cancers, or the presence of pathogenic mutations in *BRCA1/2* and/or other HRR genes in prostate cancer ^45^. Our results reveal that the HRD phenotype was not only identified across a variety of cancer types, but also associated with pathogenic variants, VUS, and wild-type status of *BRCA1/2* or other HRR genes.

Our study further expands the spectrum of molecular events that cause HRD and includes biallelic structural variants in *BRCA1/2* and other HRR pathway genes. Our results in this area confirm the data reported by Ewing *et al*., on *BRCA1/2* SVs in ovarian serous carcinomas, prostatic carcinoma, and breast carcinoma and expand our knowledge of SVs impacting other HRR genes like *RAD51B, FANCC*, and *CHEK2* among others. ^48^. We also report that cases with structural or small mutation VUS in *BRCA1/2* or other HRR genes were the most probable drivers in cases with HRD phenotype, not coinciding with other known pathogenic mutations in the HRR pathway. This suggests that those VUS are possibly related to HRD and might be considered pathogenic, supporting a previous study that reported a correlation between *BRCA1/2* VUS and the HRD phenotype ^49^.

We also interrogated those *BRCA*wt cases with HRD phenotype. These patients, identified as having HRD tumors through clinically approved testing assays like MyChoice CDx and Foundation CDx, as well as other WGS-based assays, have been reported in the literature to constitute approximately 20-30% of cases. This range aligns with the data observed in our study ^20,50,51^. Based on WGS, some of our patients had biallelic mutations in other HRD genes, like *PALB2* and *RAD51B*, known to be associated with HRD. In other cases, we identified variants in genes reported to be linked to HRR pathway dysfunction without previously known clinical impact. One example is a case with a *UIMC1* fusion in which the deletion of the AIR domain, essential for binding to the BRCA1-A complex, could have caused HRD ^36^. In some *BRCA*wt HRD cases, the genomic cause could not be uncovered by WGS, which may be due to epigenetic changes like *BRCA1/2* promoter methylation ^52-54^.

One of the main conclusions of our study is that, compared to commercial assays, WGS-based HRD assessment could clarify inconclusive or misleading results from NGS-based assays that rely on fewer signatures. Or a routine basis, we discuss such cases during a research molecular tumor board (MTB) format, a Continuing Medical Education accredited conference ^26,55-59^. Examples include WCM205 and WCM228 (detailed above), both of which showed a WGS HRD phenotype but were HRP and inconclusive by MyChoice CDx respectively. For WCM205, following a recent disease recurrence of HGSOC, the consensus was to consider PARPi as a valid treatment option, especially in light of a previous response to PBCT. For WCM228, another HGSOC with WGS HRD phenotype, and inconclusive HRD results from the commercial assay. WGS identified a biallelic structural variant in *RAD51B* as a probable cause of the HRD phenotype, rather than a monoallelic *BRCA*2 VUS seen on clinical targeted sequencing. The consensus was to continue the patient on maintenance PARPi treatment started earlier, and to which the patient was responding.

In summary, we illustrate how a WGS HRD classifier could be universally applicable in precision oncology, unearthing the HRD phenotype that may be invisible by other methods. Future functional studies will help elucidate the role of structural variants in causing HRD. We acknowledge that larger clinical response data is required to definitively assess accuracy of HRD predictions against treatment response. This study provides major impetus to evaluate WGS as a potential clinical diagnostic tool to target PARPi and PBCT responses and paves the way for both clinical validity studies and drug trial designs that include WGS methodology in their endpoints.

## Figure legends

**Fig S1.**
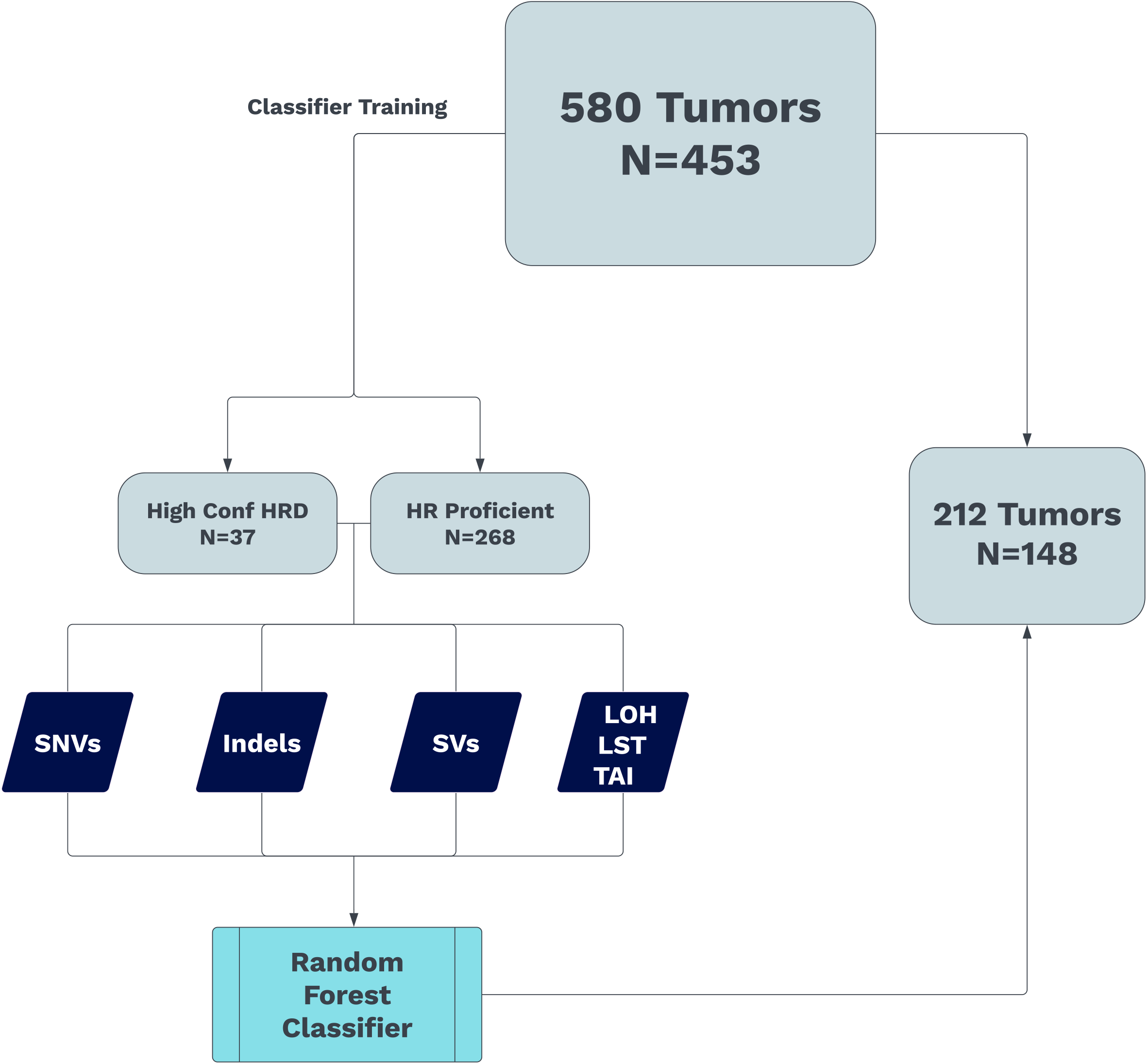
Isabl HRD Training. Training methodology described further in Hadi et al ^30^. Indels, insertions deletions; LOH, loss of heterozygosity; LST, large-scale transition; SNV, single nucleotide variant; SV, structural variant; TAI, telomeric allelic imbalance.

**Fig S2.**
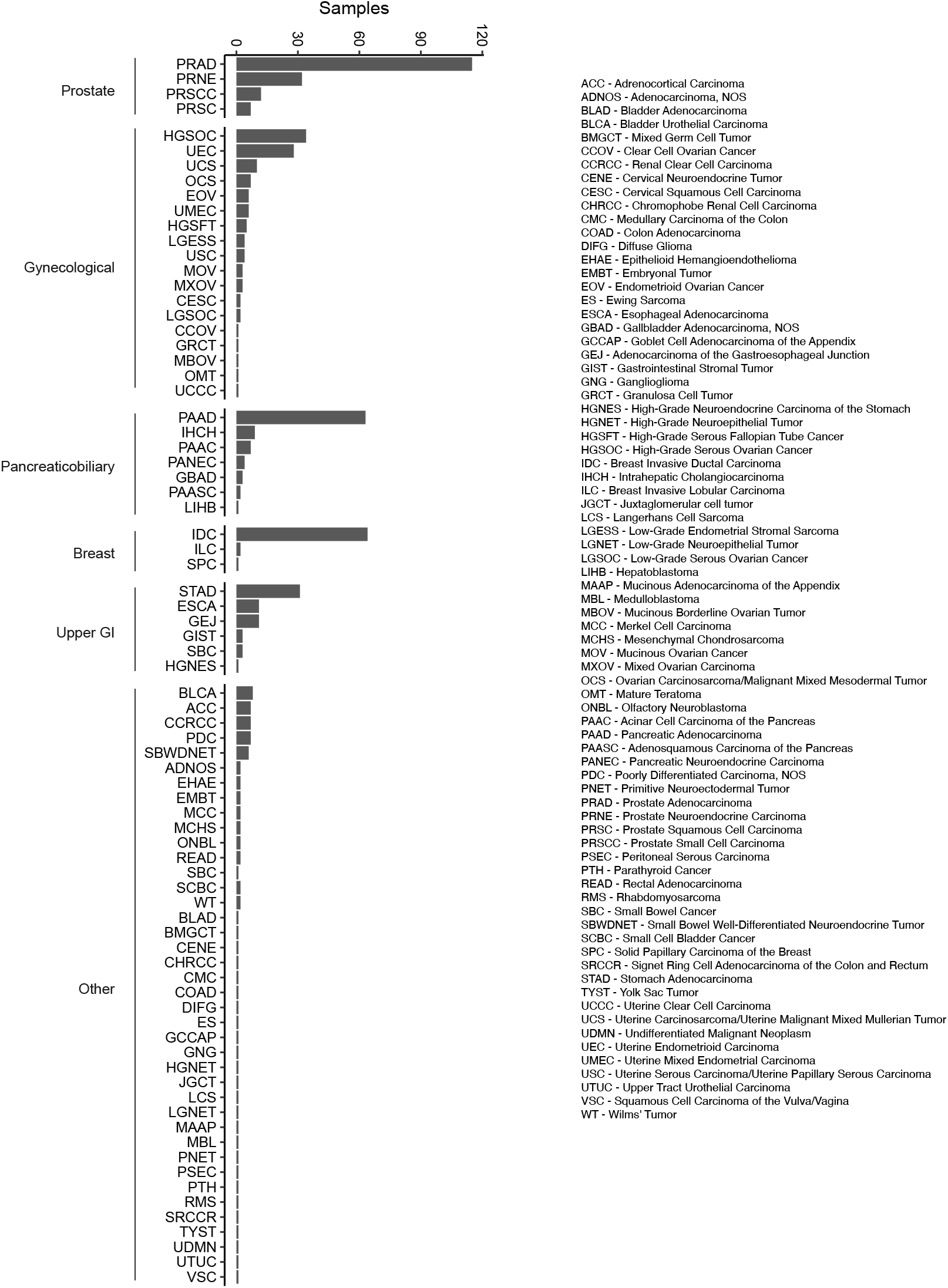
Cohort tumor subtypes.

## Acknowledgements

This work was supported by the Englander Institute for Precision Medicine. Whole-genome sequencing was performed at the New York Genome Center, supported by an agreement with Illumina, Inc., and Weill Cornell Medicine. Project support for this research was also provided in part by the Center for Translational Pathology from the Department of Pathology and Laboratory Medicine at Weill Cornell Medicine. The authors thank Ahmed G Elsaeed for project support with data extraction.

## Ethics approval and informed consent

The use of the clinical data and samples for this study was approved by Weill Cornell Medicine institutional review board (IRB) protocols # 1305013903 (Research for Precision Medicine) and # 1007011157 (Comprehensive Cancer Characterization by Genomic and Transcriptomic Profiling).

## Author contributions

M.A., K.H. and JM.M. performed study concept and design, and wrote the manuscript. JM.M, M.A., M.S., and J.M. generated the patient cohort. XX performed statistical analysis and interpretation of data. K.H., M.L., G.G., J.S.M, and E.P. performed genomic analyses. JM.M and M.A. performed the histopathological assessment. A.S. performed the FISH experiments. M.A, A.S., and K.H. prepared the figures. All authors read and approved the final paper.

## Data availability

The analyzed molecular data is available in the supplementary documents. Raw data are available upon reasonable request from the corresponding authors.

## Supplementary information

Figure S1, Figure S2

